# Leveraging Telemedicine to Improve Mnch Uptake in Kenya: A Community-Based Hybrid Model

**DOI:** 10.1101/2025.05.04.25326958

**Authors:** Edna Anab, Tabither Gitau, Erick Yegon, Nzomo Mwita, Marlyn Ochieng, Alice Koimur, Rhonnie Omondi, Stephen Smith, Harriet Andrews, David Oluoch, Rosebella Amihanda, Moses Lwanda, Erina Makhulo, Godfrey Sakwa, Phanice Akinyi

## Abstract

**Background:** Kenya faces significant challenges in providing adequate access to maternal, newborn, and child health services, particularly in remote and underserved areas. Limited infrastructure, healthcare worker shortages, and financial constraints hinder access to timely, essential care. As health systems continue to face increasing demands, Telehealth solutions offer a promising approach to bridging geographical gaps and improving access to timely and essential healthcare services. By leveraging technology, telehealth can connect patients in remote areas with healthcare providers, enabling virtual consultations, remote monitoring, and timely interventions.

**Aim:** This study evaluated the “Better Data for Better Decisions: Telehealth” initiative, funded by The Children’s Investment Fund Foundation (CIFF) and implemented by <u>Living Goods</u> and in partnership with <u>Health X Africa</u>. The innovation aimed to integrate telehealth into the Community Health Promoter framework to improve MNCH outcomes, focusing on antenatal and postnatal care. The specific objectives included increasing uptake of antenatal and postnatal care, improving the efficiency of primary healthcare delivery, and influencing relevant policies.

**Setting:** The study was conducted in Teso North, Busia County, Kenya, targeting ten community health units

**Method:** A mixed-methods quasi-experimental design was employed, incorporating key informant interviews, focus group discussions, and routine health record reviews. Data collection involved desk reviews, field data collection, and virtual data collection across three phases.

**Result:** The project exceeded its registration targets, enrolling 388 households and 551 clients. Of the registered clients, 50% engaged in consultations with Health X doctors via the hotline, which emerged as the most preferred service channel, used by approximately 88% of Telehealth platform users. The intervention positively impacted the frequency of postnatal care (PNC) touchpoints and identified at-risk women based on nutritional indicators. The average number of PNC visits within six weeks postpartum was significantly higher in the intervention sites (mean: 4.99 visits) compared to control units (mean: 3.96 visits; p = 0.003). The big wins for impact were identifying and escalating care, including completion of referrals for dangers signed in newborns, supporting positive behaviour change and improving access to clinical care in the last mile.

**Conclusion:** Integrating telemedicine into the CHW framework shows promise for improving access to and engagement with postnatal care services in underserved areas of Kenya. The hybrid model, combining virtual consultations with community-based CHW support, effectively leveraged technology and existing health infrastructure. Further research is needed to assess the impact on healthcare efficiency and policy influence fully. These findings present a compelling case for policymakers to scale telehealth as a core element of Kenya’s MNCH strategy. Part of the work led to supporting the MOH in developing Telemedicine Policy and Guidelines for Kenya.

## INTRODUCTION

Kenya, like many low- and middle-income countries, faces significant challenges in achieving optimal maternal, newborn, and child health outcomes. A key barrier is limited access to quality healthcare services, particularly in remote and underserved areas. This is further compounded by factors such as inadequate infrastructure, shortages of healthcare professionals, and financial constraints. Traditional healthcare delivery models often struggle to effectively reach these marginalized populations, resulting in disparities in health outcomes. Telehealth has emerged as a transformative tool in healthcare, offering innovative solutions to bridge access gaps, particularly in low-resource settings ^[1]^. The COVID-19 pandemic accelerated the adoption of digital health solutions, including telemedicine, as physical distancing measures disrupted traditional service delivery ^[2]^. According to the 2022 Kenya Demographic and Health Survey (KDHS) ^[3]^, the neonatal mortality rate (NMR) has declined to 21 deaths per 1,000 live births, down from 31 in 2014. Similarly, the infant mortality rate (IMR) decreased to 32 deaths per 1,000 live births from 39 in 2014. Maternal and neonatal health outcomes in Kenya remain a public health priority, with the country facing high maternal mortality rates and only modest declines in neonatal deaths. These challenges are particularly acute in underserved regions, where traditional care models struggle to reach vulnerable populations. The first 1,000 days of life represent a critical window for intervention, yet many mothers and newborns continue to lack timely, quality care, especially during the postnatal period.

Subsequently, opportunities to improve maternal and newborn survival and health through access to quality healthcare, supplemented by virtual support, during the post-partum period are underutilized and relatively unexplored, according to Living Good’s landscaping. To address these gaps, Living Goods (LG) and Children’s Investment Fund Foundation (CIFF) piloted a telemedicine solution using Community Health Workers (CHWs) to connect communities to Maternal and Newborn and Child Health (MNCH) care and services. Thus, the study evaluated the impact of this hybrid physical and telemedicine service on maternal, newborn, and child health outcomes in Malaba, Busia County, Kenya. This research is particularly important given the persistent challenges in MNCH, the potential of telemedicine to address these challenges, and the need for evidence-based strategies to improve healthcare delivery in underserved communities. By evaluating the feasibility, impact, and sustainability of this approach, the research contributes critical evidence to inform the scale-up of telehealth interventions in primary healthcare systems. It underscores the untapped potential of digital tools to support risk-stratified, patient-centered care in fragile settings—aligning with broader efforts to achieve Sustainable Development Goal (SDG) targets and reduce preventable maternal and neonatal deaths.

## METHODOLOGY

### Study Design

This study employed a quasi-experimental design with a mixed methods approach to evaluate the impact of the intervention. Figure 1 visually represents the study design. It illustrates the selection of CUs from the larger population and their assignment to intervention and comparison groups. The selection of CUs was purposively done in consultation with the Department of Health in Busia County to ensure representativeness and feasibility. The matching process is based on size, population characteristics, workforce availability, and health facility spread to minimize pre-existing differences that could confound the results. Table 1 highlights the digital tools and outcome tools utilized in the study.

**Figure 1:**
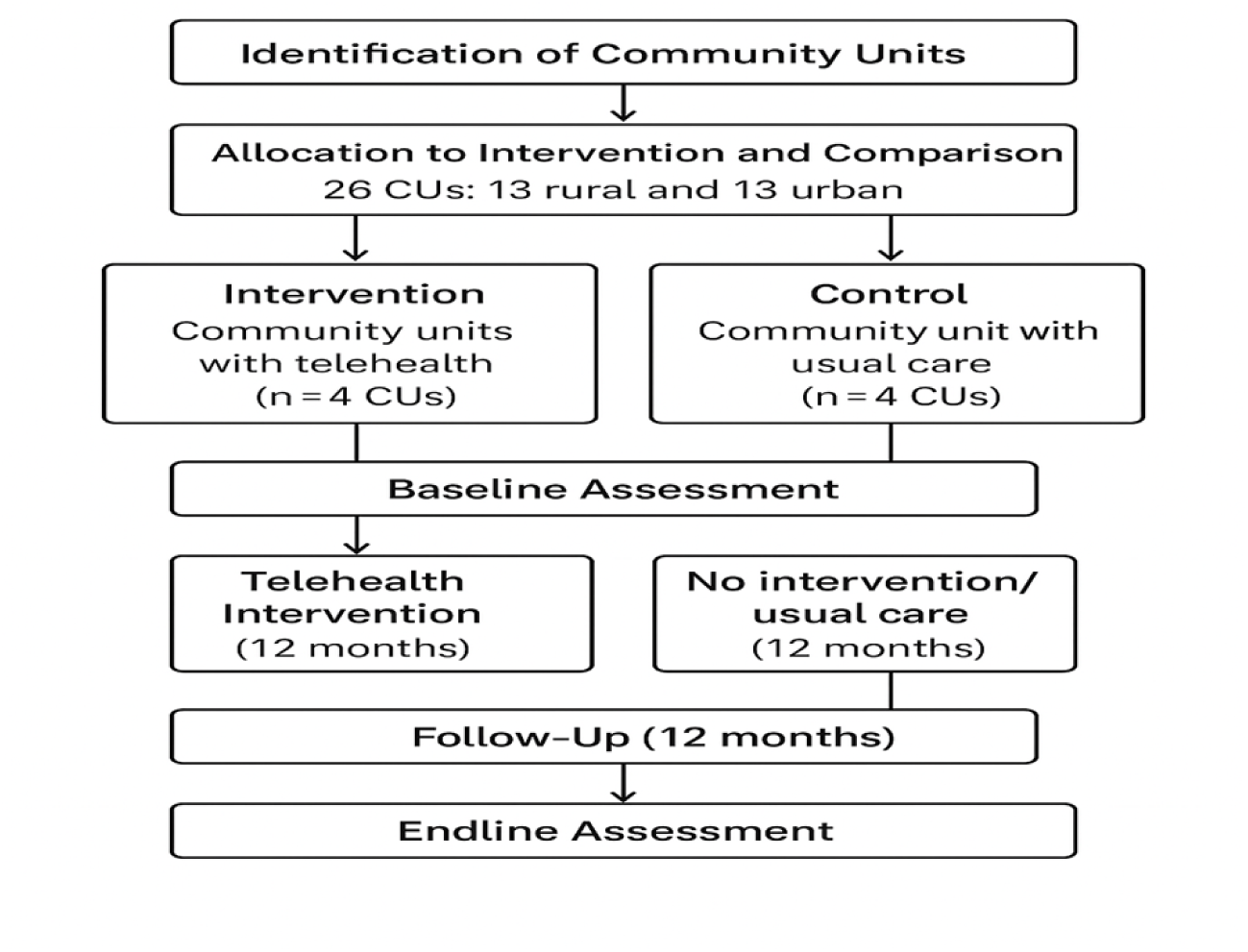
Description of the study design (quasi-experimental design)

**Table 1:**
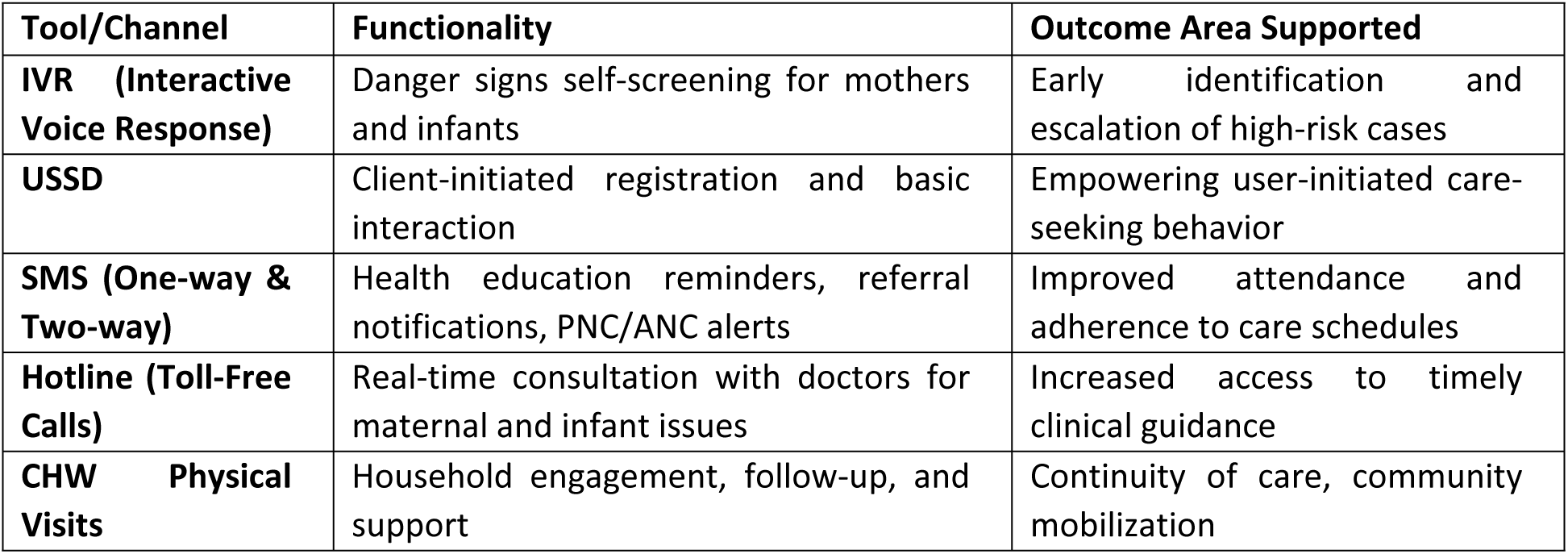
Digital Tools and Outcome Mapping.

| Tool/Channel | Functionality | Outcome Area Supported |
| --- | --- | --- |
| **IVR (Interactive Voice Response)** | Danger signs self-screening for mothers and infants | Early identification and escalation of high-risk cases |
| **USSD** | Client-initiated registration and basic interaction | Empowering user-initiated care-seeking behavior |
| **SMS (One-way & Two-way)** | Health education reminders, referral notifications, PNC/ANC alerts | Improved attendance and adherence to care schedules |
| **Hotline (Toll-Free Calls)** | Real-time consultation with doctors for maternal and infant issues | Increased access to timely clinical guidance |
| **CHW Physical Visits** | Household engagement, follow-up, and support | Continuity of care, community mobilization |

### Ethical Approval and Consent

During project implementation, all participants provided informed consent, either verbally or in writing, depending on literacy levels. CHWs were trained on ethical practices, including data confidentiality and consent procedures. For minors aged 15–17 involved in the intervention (as eligibility was later expanded), assent was obtained alongside guardian consent in accordance with ethical guidelines. Additionally, study ethical approval was sought from the JKUAT ethical review committee.

### Study Setting

The study was conducted in Teso North Sub County, Busia County, Kenya. Teso North was chosen due to the existing healthcare access challenges, especially concerning antenatal and postnatal care. The evaluation focused on 10 Community Health Units (CHUs) within Teso North. Six of these CHUs (3 peri-urban and 3 urban) were assigned to the intervention group, while 4 served as the comparison group. As shown in Figure 1, ten community units were selected from a total of 26 in Teso North, Busia County, Kenya. Six of these CUs were assigned to the intervention group, while the remaining four served as the comparison group.

### Study Population and Sampling

The study population consisted of households within the purposively selected Community Units (CUs) in Teso North, Busia County, Kenya, that had children under 6 weeks old and/or pregnant women of reproductive age (18-49 years). All eligible households within the chosen CUs were included in the study, forming the total sampling frame.

A cluster sampling approach was used, where the CUs represented the clusters, and all eligible households within those clusters were included. This targeted sampling ensured the evaluation focused on the population most relevant to postnatal care interventions. The study included 129 surveys completed by mothers (20 telehealth service non-users and 109 users of the intervention) and seven FGDs with mothers. Additional FGDs were conducted with CHWs and local government staff to gain broader perspectives on the intervention’s impact.

**Table 2:** Telehealth Intervention Timeline and Rollout Phases.

| Phase | Timeline | Key Activities | CHUs Involved | Tools/Channels Deployed |
| --- | --- | --- | --- | --- |
| **Phase 1: Design & Co-Design** | Feb – July 2023 | Formative research, feasibility assessments, partner selection (Health X Africa), design approvals | N/A | Planning only |
| **Phase 2: Pilot Rollout – Phase I** | June – Oct 2023 | Launch with 17 CHWs in two CHUs (Moding and Komiriai); IVR, hotline, SMS rollout | **Moding, Komiriai** | USSD, Hotline, One-way SMS, IVR |
| **Phase 2: Pilot Rollout – Phase II** | Nov 2023 – Feb 2024 | Expansion to 22 CHWs in two more CHUs; wider IVR rollout, expanded eligibility (15+ years) | **Ikapolok, Onyunyur** | Expanded IVR, Hotline, Personalized SMS |
| **Phase 3: Optimization & Scale** | Mar – Sep 2024 | Two-way SMS added, standardized SMS schedule; evaluation of engagement, referrals, and usage | All 4 pilot CHUs | Two-way SMS, Standardized Messaging |
| **Endline Evaluation** | Sep 2024 | Final data collection, analysis of uptake, impact, and referral completion | All Intervention Sites | Monitoring tools, clinical data, FGDs & KIIs |

### Intervention Description

The project aimed to design and implement an integrated virtual and physical 24-hour triage solution, allowing clients to self-screen and be directed towards appropriate care pathways. The goal was to enhance care-seeking behaviours, reduce delays in care, and improve efficiency in delivering care.

This intervention integrated a telemedicine solution into an existing community health platform, with the goals of:

- Improving the uptake of essential healthcare services, specifically postnatal care.
- Increasing the efficiency of primary healthcare delivery by reducing the need for in-person visits.
- Ultimately, influencing national and county-level policies to create a supportive environment for telemedicine innovations.

The intervention incorporated Interactive Voice Response (IVR) for self-screening of danger signs for mothers and infants, contributing to early identification of danger signs and escalation for better health outcomes. Additional SMS for reminders and notification of referrals from Health X to the CHWs and supervisors. It operated alongside the routine service delivery model of CHWs, offering an additional layer of support and access to care. This hybrid approach leveraged both technology and existing community health infrastructure to address challenges in accessing postnatal care services (Refer to Table 3).

**Table 3:** Registered clients and adoption rate by Community Unit.

| Telehealth Adoption Rate by Community Unit |  |  |  |  |
| --- | --- | --- | --- | --- |
|  | CU Description | Women | Infants | Duration of Implementation |
| Moding | Rural | 52/136 (38%) | 34/61 (56%) | Phase 1: June '23 |
| Komiriai | Urban | 59/114 (52%) | 27/53 (51%) | Phase 1: June '23 |
| Ikapolok | Peri-urban | 44/73 (60%) | 7/30 (23%) | Phase 2: Nov '23 |
| Onyunyur | Rural | 34/61 (56%) | 11/18 (61%) | Phase 2: Nov '23 |
| Non-users |  | 178 | 76 |  |
| Missing |  | 21 | 8 |  |
| **Total** |  | **388** | **163** |  |

#### Study Phases

The project had a 24-month timeline divided into three phases: Exploration & Co-Design (6 months); Prototyping and Early Testing (6 months), and Pilot Implementation & Evaluation (12 months). The evaluation process was structured into three distinct phases:

**Phase One**: *Design Phase: -* This phase entailed formative research, sector landscaping, and early-stage exploratory research on compatibility and feasibility of modalities. Additionally, this phase entailed scoping and selection of a partner in technology and telemedicine service provision, settling on HealthX Africa.

**Phase Two**: *Innovation Implementation:* - This phase entailed submission of project design documents to the donor (CIFF) for sign off and seeking ethical approvals. This was followed by the launch of Phase I of the intervention, where 17 CHWs in the first two Community Units (Moding and Komiriai in June 2023, with review and design pivots. The pilot went live with the USSD (English & Swahili), hotline, and SMS channels. Phase II, spanning from November 2023 to February 2024, engaged an additional 22 CHWs from Ikapolok and Onyunyur Community Health Units. In terms of the intervention, this phase included the full deployment of IVR, extending eligibility to include clients aged 15 years and above. Phase III began in March to September 2024 with design pivots where the Two-way SMS was launched in addition to personalized SMS and a standardised schedule of health information SMS.

**Phase Three:** *Endline Evaluation: -* The endline evaluation was conducted in September 2024.

Figure 2 shows the telehealth engagement journey from community mobilization and registration to postnatal period.

**Figure 2:**
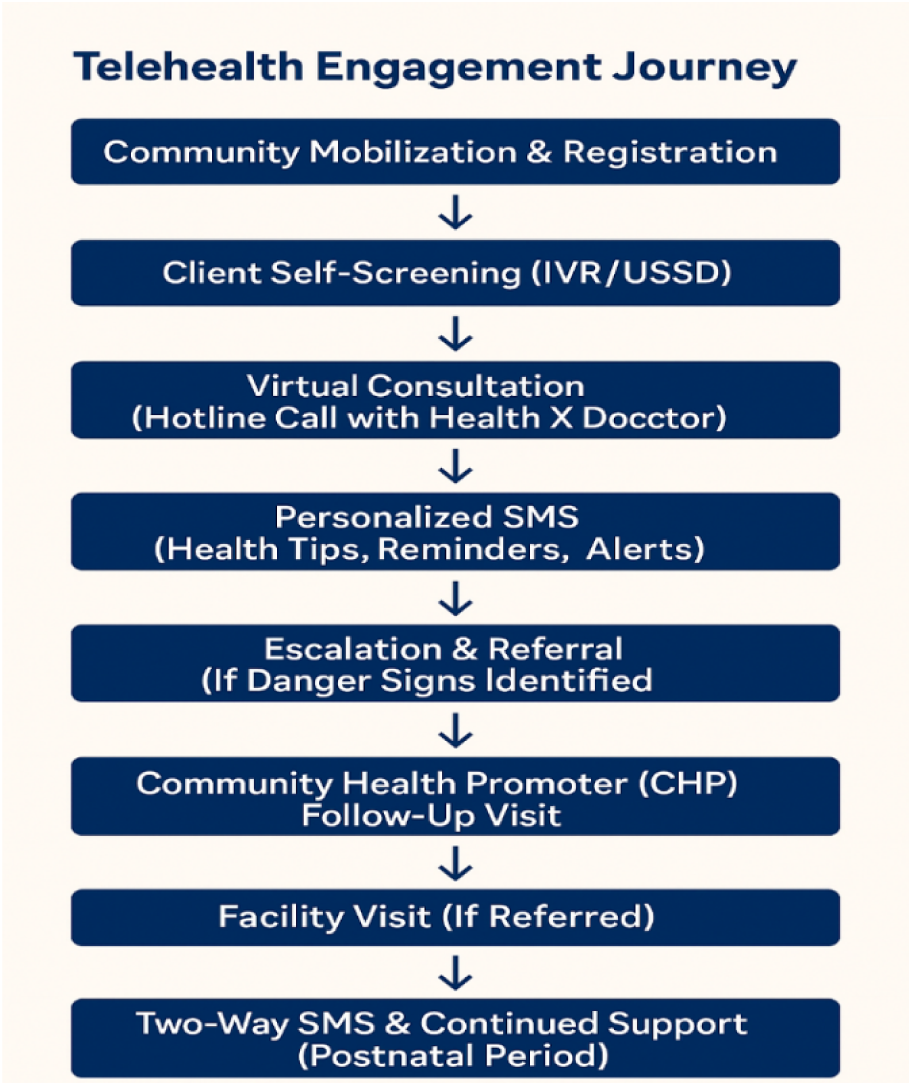
Telehealth Engagement Journey

### Data Collection

The data collection for this study employed a mixed-methods approach, gathering both quantitative and qualitative data across three distinct phases:

**Desk Review:** This phase was conducted between July 15th and September 6th, 2020, and involved reviewing project documents, Health X databases, and Local Government databases. The purpose of this review was to establish background and context for the study, as well as to triangulate findings from the primary data collection activities. The desk review culminated in an inception report outlining the finalized evaluation methodology, timelines, and data collection tools.

**Field & Virtual Data Collection:** This phase took place from September 9th to 23rd, 2024, utilizing a stratified, criteria-based, purposive sampling method to select participants. Several data collection methods were employed:

**Key Informant Interviews:** Semi-structured interviews were conducted with various stakeholders, including county and sub-county health officers, mothers participating in the intervention, a local government telehealth champion, 4 County and Sub County Ministry of Health staff, nurses from referral facilities, and Health X staff. Tailored interview guides were developed for each stakeholder group to ensure relevant and focused data collection.

**Focus Group Discussions:** A total of 11 FGDs were held, involving 168 primary beneficiaries (ANC and PNC mothers), and 38 Community Health Workers. Participants were segmented by telehealth service user engagement levels (high, medium, and low) for both mothers and CHWs.

**Monitoring Program Data**: Project performance indicators related to service utilization and health outcomes were analyzed using data spanning from July 15th to September 30th, 2024. This quantitative data was extracted from project data.

### Data Analysis

Both quantitative and qualitative data analysis techniques were applied. Quantitative data were analyzed using Stata® version 15, employing descriptive and exploratory statistics. For qualitative data, the audio recordings were transcribed and imported into Dedoose for coding and further analysis. Analysis across all transcripts was conducted using a constant comparative method to identify emerging themes and their repetitions and variations.

## RESULTS

This section summarizes the key findings on the hybrid virtual telehealth and Community Health Worker intervention in Teso North, Kenya, focusing on user registration, adoption, CHW Influence in the intervention uptake, service uptake, impact, nutrition, and sustainability.

### User Registration and client profile

The hybrid model and virtual platform supported 388 households, reaching 551 clients (388 mothers and 163 infants) through 39 Community Health Providers. Program data indicated that the majority of the registered mothers were young adults aged between 20 to 29 years at 59% (n=228), with the majority enrolling during their second trimester at 27% (106 women). 113/ 129 (88%) of registered women surveyed were married, while 12% (16) self-reported as single mothers. Regarding education, more than half of the mothers had attained primary and secondary education at 62(48%) and 50(39%), respectively. By the end-line study in September 2024, 77% (99/129) had delivered. Among those surveyed, 88% (113/ 129) were married and 12% (16) were single mothers, with most having primary, 48% (62), or secondary, 39% (50) education. It was critical to note that almost half of these mothers had no source of income at 48% (n=62), translating to 50% of the mothers reporting to have no independent income whatsoever (n=65). By the end-line study in September 2024, a significant majority (77%, 99 women) had given birth, while 60 eligible women (either pregnant or within six weeks postpartum) remained enrolled at the time of reporting. These results highlight telehealth’s potential to engage women earlier in their pregnancy, despite the project’s primary focus being on postnatal care.

### Adopters: Client profile and frequency of use

Almost half of the registered women and the same proportion of infants utilized the Telehealth services. High adoption of Telehealth services was noted in Moding and Komiriai CUs, which were the Phase one intervention sites. While the two phase two sites, almost at par with about 15% contribution each to the total adoption rates.

An analysis focusing on women’s engagement with Health X measured through the frequency of clinical calls to Health X by the mothers categorized as follows: High, medium, low and none characterized by 4 times or more, 2-3 times, one and no engagement with the Health X doctors after registration, respectively. This engagement could have been initiated via text that would lead to a follow-up call or a direct call to Health X. High adoption was noted in Moding (Phase 1) and Komiriai (Phase 2) CUs, both accounting for 58% (111/189) of all adopters (Table 4).

**Table 4:** Frequency of use by community unity.

| Frequency of use by community unity (Jun 2023- Sep 2024) |  |  |  |  |  |  |
| --- | --- | --- | --- | --- | --- | --- |
| Community Unit | High | Medium | Low | Non-users | Total | Total Users |
| Moding | 11(3%) | 19(5%) | 22(6%) | 69(18%) | 121(33%) | 52(13%) |
| Komiriai | 3(1%) | 26(7%) | 30(8%) | 51(13%) | 110(30%) | 59(15%) |
| Ikapolok | 8(2%) | 22(6%) | 14(4%) | 33(9%) | 77(21%) | 44(11%) |
| Onyunyur | 6(2%) | 12(3%) | 16(4%) | 25(6%) | 59(16%) | 34(9%) |
| Missing Data |  |  |  |  | 21(5%) |  |
|  | 28(8%) | 79(22%) | 82(22%) | 178(49%) | 100% | 189(49%) |

**Table 5:**
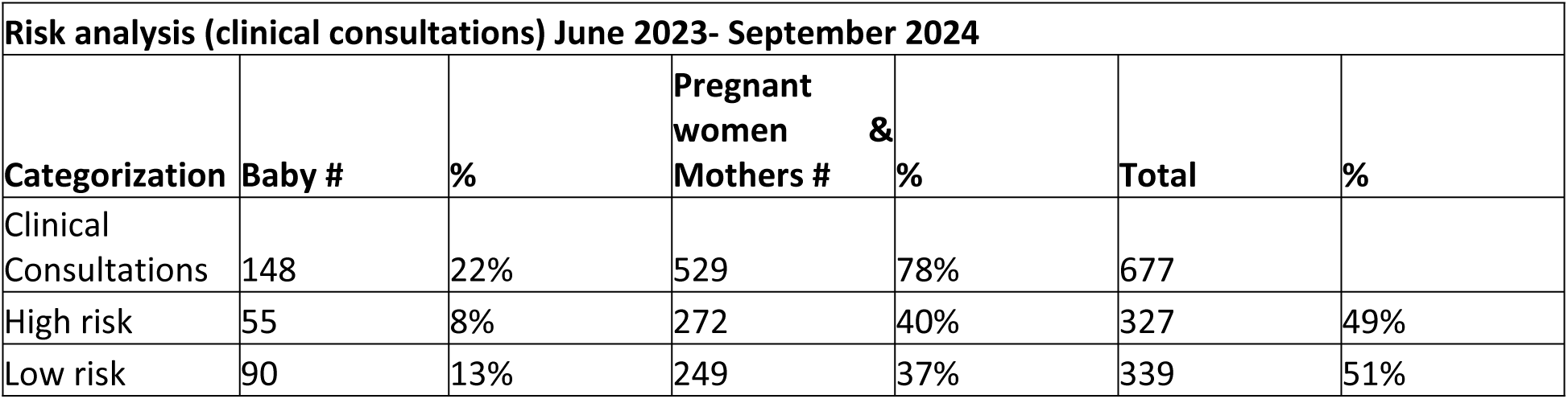
Clinical Consultations Risk analysis.

| **Risk analysis (clinical consultations) June 2023- September 2024** |  |  |  |  |  |  |
| --- | --- | --- | --- | --- | --- | --- |
| **Categorization** | **Baby #** | **%** | **Pregnant women & Mothers #** | **%** | **Total** | **%** |
| Clinical Consultations | 148 | 22% | 529 | 78% | 677 |  |
| High risk | 55 | 8% | 272 | 40% | 327 | 49% |
| Low risk | 90 | 13% | 249 | 37% | 339 | 51% |

CHWs observed that high adopters of the virtual health service were typically individuals with a high perceived need, such as first-time mothers, adolescents, or those with a history of obstetric complications. They also tended to have slightly higher literacy and education levels, owned phones, or had supportive partners willing to share their phones and access to the service. A key driver of adoption was the quality of virtual customer service. Many users reported receiving more personalized care, longer consultations, and access to broader medical expertise than they typically received from local health facilities or CHWs.

In contrast, non-adopters often cited limited time and low perceived value of the service, believing they would be referred to a facility for care regardless. CHWs reported that these mothers often had low literacy levels, poor initial customer service experiences (such as missed call-backs or long wait times), difficulty using the platform independently, and limited phone access—often sharing devices with CHWs, partners, or in-laws, or using inactive SIM cards. CHWs also noted that higher-income or working-class mothers were more likely to have health insurance or prefer private health services.

> *“Mostly, we would have encounters with mothers between 19 to 35 years who were willing to register more than those who are beyond 35 years. Those above 35 years old felt uncomfortable engaging the (virtual) doctors.” **FGD, CHW***

> *“For my registration, the younger pregnant women responded easily, as they don’t have pregnancy experience. They respond easily so that they can get help.” **FGD, CHW***

### Motivators and key enablers for client registration and client use

Overall, while the majority of the virtual CHWs delivered relatively well in enrolment, a strong backbone of performance management by supervisors was required to maintain momentum. CHW marketing of the virtual service primarily focused on the hotline as an emergency or clinical counselling service. Majority, 62% (24/39 CHWs) trained as virtual ambassadors onboarded a satisfactory level (10 – 28) of clients since project launch.

**Enablers:** Key drivers of successful performance include strong supportive supervision and confidence of certain CHWs to onboard and integrate registration as part of independent day-to-day activities; good levels of CHW technical proficiency (to guide households and handling and troubleshooting); and a clear understanding of roles and responsibilities. In addition, those that were successful with onboarding were overall well performing CHWs, with some of the factors that worked in favour included having a reliable means of transport which increased their reach and access, regular CHW engagements with clients to promote telehealth utilization, and CHWs focusing on key health areas such as pregnancy enabled them to perform higher than their counterparts.

During the pilot, we focused primarily on marketing channels through CHWs directly (Figure 2). However, insights from the FGDs suggest that clients who were initially resistant (mid - late adopters) to the service were influenced by peer recommendations and stories of the value of telehealth. According to our Health X KII, the use of snowballing for pregnant mothers to refer other pregnant mothers for registration led to a surge in registration.

**Figure 3:**
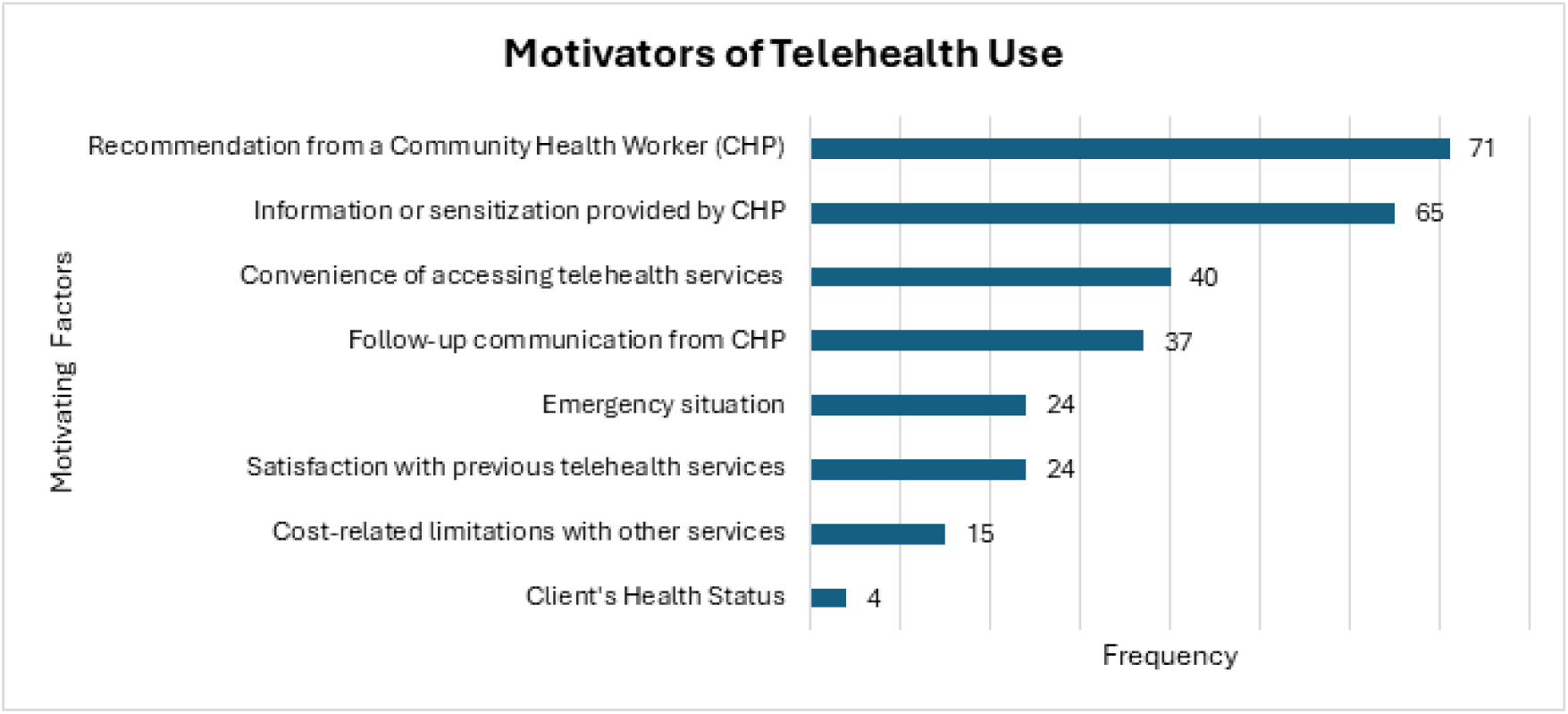
Motivation of Telehealth Use

Peer Recommendations: “*When we started the registration, most of them were unwilling. But the moment those who registered started engaging the telehealth doctors and got to see the benefits, they then came back and passed the message in the community. They became our good ambassadors, and this made our work easier. Those who are registered will go and tell the others.” FGD, CHW from Moding*

**Inhibitors:** Barriers to the registration of clients to telehealth services included misconceptions about telehealth services, spousal refusal, lack of mobile phones, technical challenges, socio-cultural aspects, and privacy concerns (i.e., disclosing a pregnancy early or HIV status). The low CHW performers reported that some clients held misconceptions about the purpose and efficacy of telehealth services, which led to hesitations in registering. In some households, women faced resistance from their spouses in utilizing telehealth services. In addition, limited access to mobile phones made it hard for them to access telehealth services.

**Figure 4:**
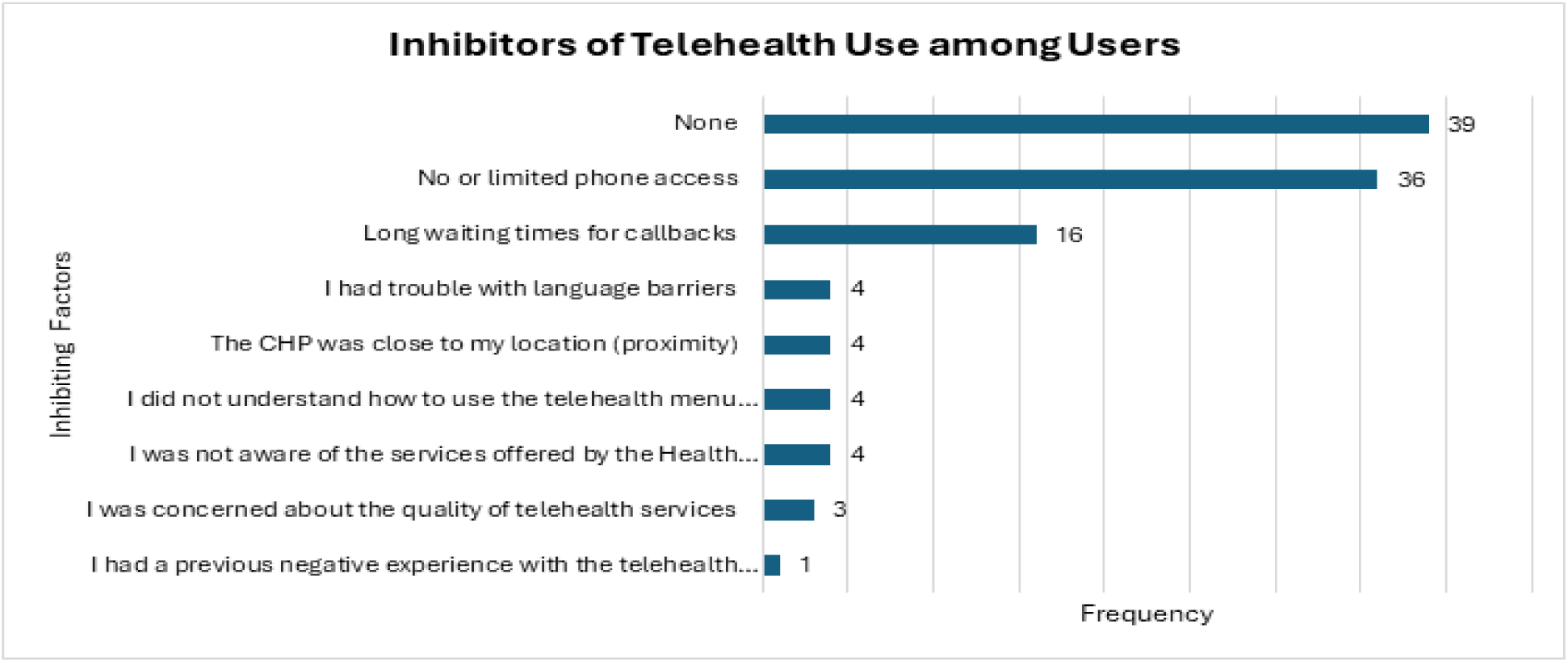
Inhibitors of Telehealth Use

### Uptake of services: functions & channels

Strong client preference for the toll-free clinical calls remains the predominant channel accessed and used by households, with very limited use of the other USSD or IVR enabled functions and features (hail your CHW, danger sign risk assessment, report a delivery). SMS reminders likely prompted positive behaviour change, with 64% (83/129) of women reporting the reminders improved their timely clinical ANC and PNC attendance. On exploring how the respondents used Telehealth. The majority of the respondents (n=90, 83%) used the platform to speak to/consult a doctor when they or their baby were ill. This was followed by 68 respondents who reported registration of themselves or their baby on delivery, while very few (n=6) used the platform to hail their CHW.

**Figure 5:**
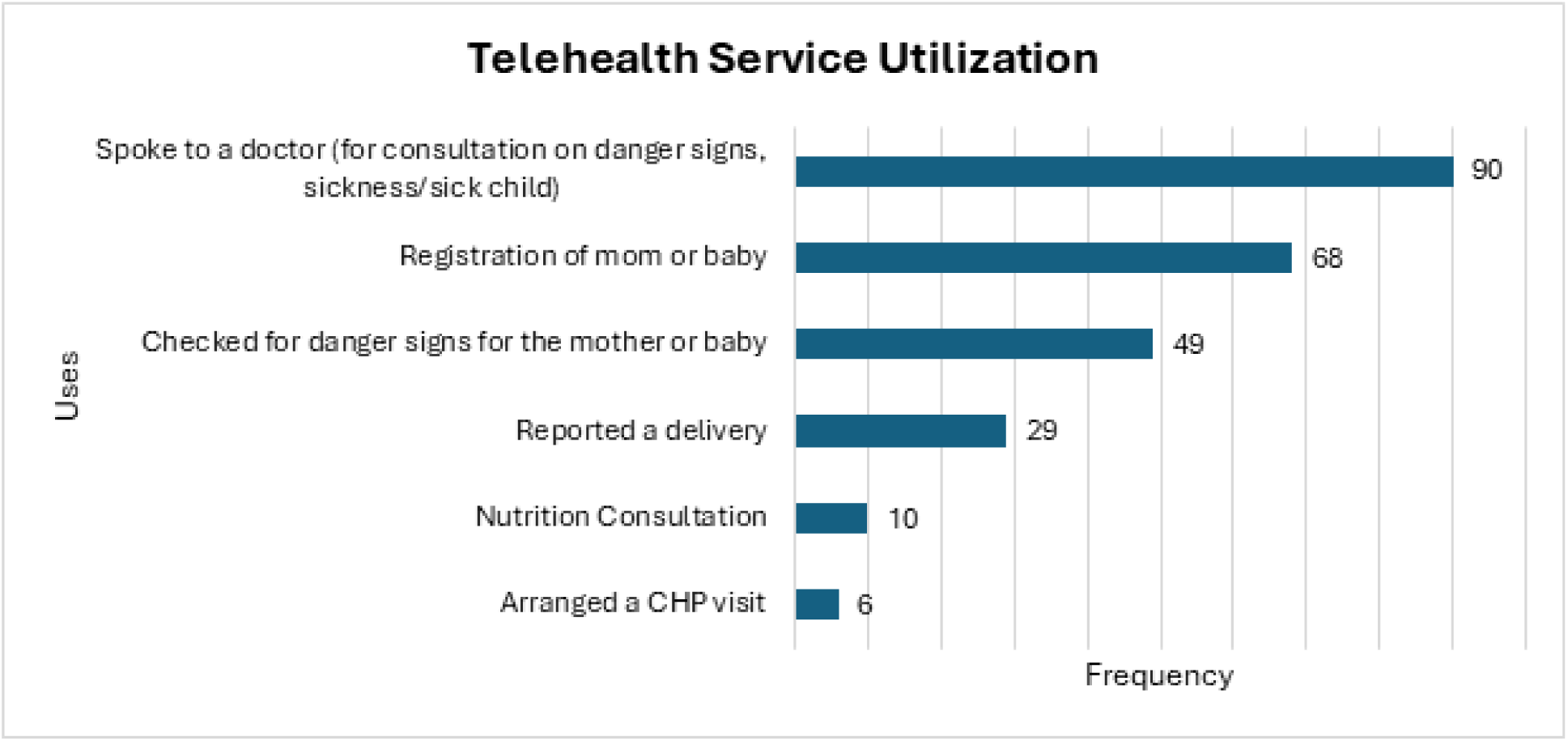
Telehealth Services Utilization

Both in IVR and USSD, the risk assessment tool (users can hear a list of danger signs associated with new mothers or infants in the PNC period) and the “Hail your CHW” had very limited uptake. In part early insights suggest this may have been due to a) low perceived value (clients already have the CHW phone number) b) a lack of active marketing by CHWs of these features c) that calling the virtual Doctor directly via the hotline was a more reassuring and direct experience for the user and d) that the virtual triage (eg danger signs) would escalate to a virtual doctor consultation).

### Telehealth Impact

#### Management of high-risk cases and danger signs

Between June 2023 and September 2024, a total of 1,108 calls were made to the hotline, of which 61% (677 calls) were clinical encounters. The majority of these clinical consultations (78%, 529/677) were related to maternal health, while 22% (148/677) concerned infant care. Telehealth was valued by users as a confidential and accessible channel for seeking advice and support, especially for maternal health needs. Most clinical calls (64%, 435/677) were effectively managed by Health X without the need for referral, while 12% (79/677) required onward referral to local health facilities or specialists. Only six cases during this period required a prescription. Notably, 23% (248) of the clinical calls occurred outside of the standard 8AM –5PM CHW working hours, highlighting the extended access offered by telehealth services. High-risk cases accounted for 49% (327/677) of all clinical consultations, with the majority (83%, 272/327) involving pregnant women or mothers, compared to 17% (55/327) involving infants (Table 3)

#### Completion of Referral

Across the project duration (July 2023 - September 2024), 92% (3,630/3,967) of referrals were completed in the intervention sites, compared to 97% (1,873/1,938) in the control site. While the control site had a higher percentage, the absolute number of completed referrals was substantially higher in the intervention sites (3,630 vs. 1,873). The report notes, ‘Based on the program data, control sites were referring to a higher number of infants with danger signs, the absolute loss to follow up in the control site was much higher at 329 infants compared to 62 infants in the intervention site’. This suggests the telehealth intervention effectively triaged cases, potentially reducing the burden on higher-level care facilities. Furthermore, the intervention sites achieved a consistently higher performance on proportion of completed referrals for infants with danger signs compared to the control site and maintained this positive trend even during periods of disruption, as shown in the figure below.

**Figure 6:**
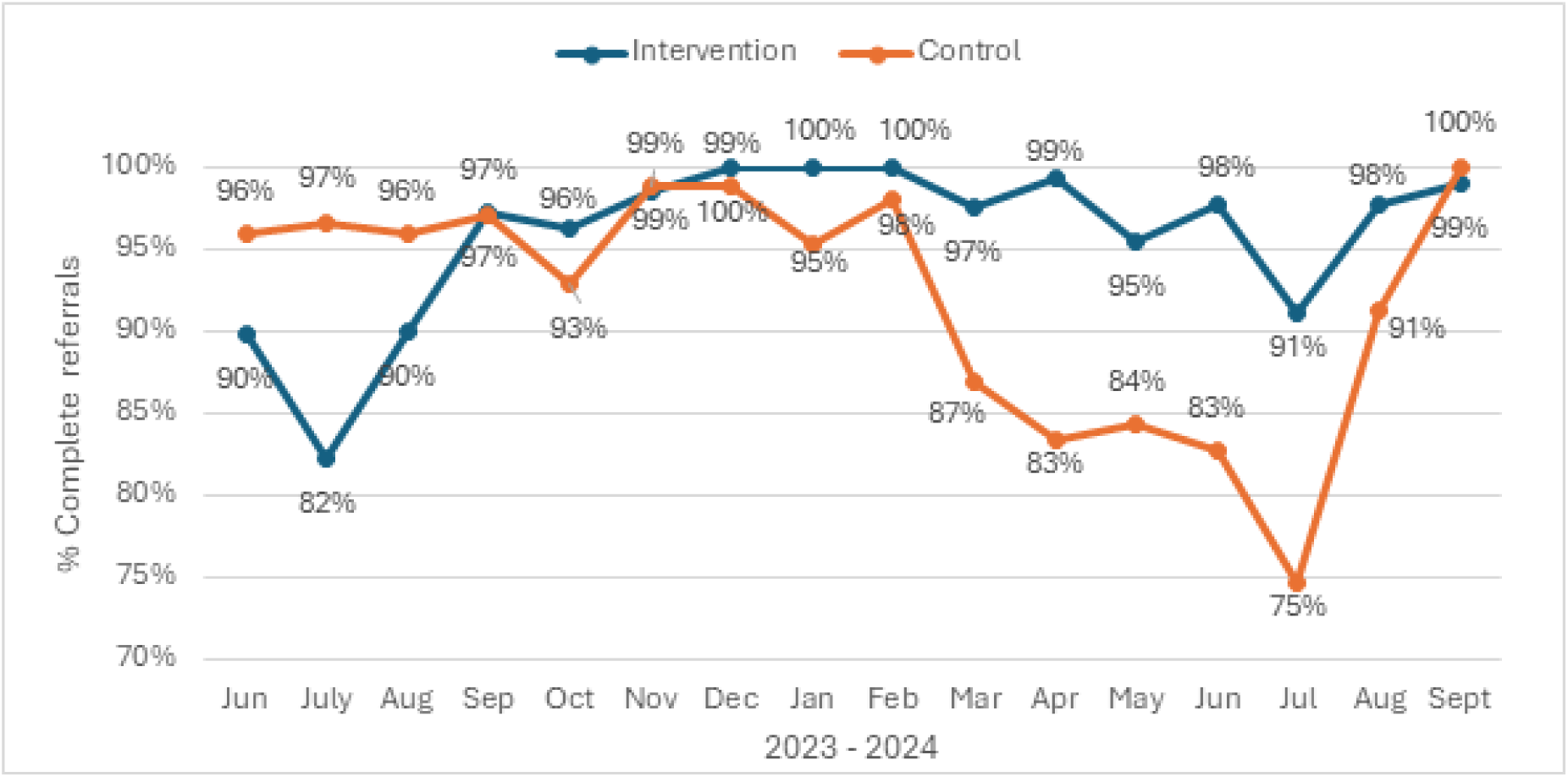
Percentage of Newborns Completing Referrals

#### Number of PNC touchpoints

The average number of PNC touchpoints between CHW and client within 6 weeks of delivery was, on average, statistically (p-value: 0.003) slightly higher in the intervention site at approximately 5 visits (4.99) compared to approximately 4 visits (3.96) in the control community units. However, these averages mask a greater monthly fluctuation of visits in the intervention sites during pilot implementation.

> *“Those women who were registered on Telehealth after delivery know the best practices on how to take care of a newborn. Those who were not registered, if the CHW has not also coached them correctly, don’t know the best practices for newborn care. They have enough education on newborn birth care practices. Those on telehealth, they talk, they speak to the doctors frequently, unlike those who are not registered, so they tend to get more knowledge than those who are not registered.” **FGD, CHW***

Survey with mothers revealed that the experiment group, on average, has a significantly higher number of PNC visits (4.99) compared to the control group (3.96) but with more variability. Hence, while the intervention in the experimental group may lead to more visits on average, the results are more variable and less consistent. Across June 2023 – September 2024, trends for the unique women/infants visited at least once by their CHW within 48 hours were on average 85% compared to the control of 87%.

Women/infants receiving all three PNC touchpoints’ on time within 6 weeks, was not positively impacted by the virtual platform when we observe the trends between intervention and control site (334/625 (53%) vs 965/1589 (61%), however due to the difference in the total number of women being served, the intervention site experienced significantly fewer cases of women/infants lost to follow-up during that period (291 compared to 625).

#### Perceived value and impact for CHWs

According to FGDs and KIIs with CHWs and Supervisors, the introduction of the hybrid virtual model resulted in several system efficiencies and benefits:

- Improved follow-up by making it easier for CHWs to monitor pregnant and post-partum mothers through notifications
- Boosted CHWs’ confidence and their standing within the community through the association with Health X doctors,
- Reducing the need for extensive education during sessions, with registered clients more likely to be receptive and knowledgeable with positive health-seeking behaviours
- Telehealth simplified referrals and disease identification, making it easier for CHWs to refer clients to health facilities, and streamlined follow-up tasks by alerting CHWs to client needs.

- Health X doctors assisted CHWs in encouraging clients to visit health facilities when referred.
- Provided a way to more immediately escalate a health concern from the household

For the highly adopting clients and the highly performing CHWs, the KIIs and FDGs revealed a level of promising synergy between client, CHW, and virtual telehealth solutions, as intended in the original design:

> *“The virtual support brought the clients closer to me during the ANC period. Each time the client engages the telehealth doctor, they would call me, and any time I received SMS about a certain client, I would visit them… I would say 90% drew closer due to the virtual support,” FGD, CHW High Adopter*.

> *“…For those (clients) who do not read their messages, I communicate with them as soon as possible and pass the message to the relevant personnel in Telehealth. This has helped to facilitate faster communication,”* FGD, CHW.

## DISCUSSION

This study evaluated the integration of telemedicine into Kenya’s CHW framework to improve MNCH outcomes. The findings highlight the critical role of CHWs in facilitating telehealth adoption, the importance of early maternal engagement, and the influence of community-driven adoption patterns on service utilization. While the intervention demonstrated significant improvements in healthcare access, sociocultural barriers, program awareness, and CHWs capacity remain a key challenge to its long-term sustainability.

### Enhanced access and utilization of maternal health services

The telehealth intervention exceeded its registration targets, enrolling 388 households and 551 clients, demonstrating strong community engagement and improved access to healthcare services. The significant proportion of women enrolling in their second trimester suggests that the platform successfully encouraged earlier engagement in maternal healthcare. Additionally, the increase in PNC touchpoints indicates a positive shift toward more consistent maternal and neonatal healthcare utilization. This is in line with findings from a retrospective that was done to evaluate the impact of telemedicine, the study found an increase in PNC attendance and postpartum depression screening ^[4]^ (Arias et al. 2022) The high reliance on hotline consultations as the preferred communication channel further reinforces the feasibility of integrating telehealth into community health service delivery.

### Improved referral completion and follow-up

The intervention achieved a high referral completion rate (92%), with more referrals completed at intervention sites than control sites (3,630 vs. 1,873). Although control sites had a slightly higher completion percentage (97% vs. 92%), they also had a higher number of missed referrals (329 vs. 62), highlighting telehealth’s effectiveness in triaging cases and ensuring timely follow-up. This likely reduced the burden on health facilities and improved continuity of care. These findings align with previous research showing that telehealth can enhance adherence to care during prenatal and postnatal periods while complementing essential in-person services ^[5]^. SMS reminders also played a key role in boosting appointment adherence, emphasizing the value of mHealth strategies in promoting patient engagement and care-seeking behavior, consistent with findings from ^[6]^ Knop et al. (2024), where SMS increased and timely child immunization.

### Effectiveness of the hybrid telehealth model

The study highlights the success of a hybrid model that integrates virtual consultations with community-based support from CHWs. This approach facilitated community buy-in, strengthened trust, and ensured continuity of care. By leveraging CHWs as intermediaries, the intervention bridged digital literacy gaps and enhanced service uptake. However, CHWs faced initial challenges in integrating telehealth into their workflows, emphasizing the need for ongoing capacity-building, technical support, and structured supervision to maximize their effectiveness ^[7]^. It aligns with broader evidence showing that telemedicine can improve maternal outcomes in Africa but faces challenges such as infrastructure and digital literacy gaps ^[8]^.

### Addressing healthcare access barriers

The telehealth initiative effectively tackled healthcare access challenges in remote and underserved communities. The ability to consult healthcare providers via mobile phones reduced the need for physical travel, addressing both cost and distance barriers. However, sociocultural factors such as spousal approval and limited program awareness impacted enrolment rates, particularly among certain demographics. The study also identified gaps in phone access and network connectivity, which need to be addressed to ensure equitable access to telehealth services. This study supports the growing evidence that adopting telehealth technologies can enhance the antenatal care experience for women and improve access, while potentially reducing healthcare costs without negatively affecting maternal or neonatal health outcomes. However, as highlighted in recent literature (e.g., *Telehealth in antenatal care: recent insights and advances*), further research is needed to assess the impact of telehealth on rare but critical outcomes such as maternal and neonatal mortality ^[9]^. Additionally, the study echoes concern from ^[10]^ Berkley et al. (2024), which highlight that many high-risk children go undetected while low-risk children often receive unnecessarily intensive care. This imbalance underscores the importance of integrating telehealth solutions to ensure timely identification and escalation of high-risk cases, ultimately optimizing healthcare resource allocation and improving survival outcomes.

## LESSONS LEARNT

- **Leverage CHWs as central catalysts**: CHWs with strong technical skills, community trust, and supportive supervision played a pivotal role in driving adoption and engagement with telehealth. Future rollouts should prioritize training, motivation, and digital literacy support for CHWs.
- **Start with strong virtual customer service**: Early user experiences with the hotline significantly influenced platform perception. Timely callbacks, respectful tone, and continuity of care should be non-negotiable standards.
- **Target first-time and high-risk mothers**: These groups showed the highest uptake, indicating a natural demand for personalized, remote support. Tailoring messaging and outreach toward them can boost adoption.
- **Enable peer-to-peer Influence**: Snowball referrals and positive word-of-mouth among mothers were powerful adoption drivers. Incorporate structured community advocacy and testimonial-based messaging into future strategies.
- **Simplify tech tools and messaging**: Features like “Hail your CHW” or IVR self-screenings saw low uptake due to complexity or poor awareness. Ensure user-centered design, ongoing promotion, and clarity of value in future rollouts.
- **Address device and access barriers**: Limited phone access and shared devices were major barriers, especially for adolescent or low-income mothers. Future designs should explore shared device protocols, offline functionality, and equity strategies.
- **Avoid overreliance on one channel**: The hotline dominated usage, while other channels (e.g., USSD, IVR) underperformed. Balance promotion efforts across channels to maximize the platform’s full potential.
- **Plan for sociocultural resistance**: Factors like spousal refusal, privacy concerns, and stigma delay uptake. Proactive community engagement, male partner sensitization, and anonymous or discreet features should be embedded in design.

## RECOMMENDATIONS FOR FUTURE IMPLEMENTATION

To enhance the impact and sustainability of telehealth interventions, the study recommends:

1. **Optimize a Digitized Closed Loop Referral System (CLR):** Develop an integrated digital platform that connects all key stakeholders, CHWs, supervisors, telehealth providers, and health facilities enabling seamless coordination and follow-up.
2. **Ensure Interoperable Client Data:** Integrate interoperable client records and data flows into the national electronic Community Health Information System (eCHIS) to support a comprehensive, data-driven approach for identifying and managing high-risk women and infants.
3. **Use Data for Risk Identification & Stratification:** Pilot and refine risk stratification models using client data, including testing predictive algorithms and applying both low-tech and AI-enabled diagnostics to identify high-risk women and children under two years of age.
4. **Deliver Personalized & Precision Care at the Household Level:** Enrol clients into customized care pathways based on their health profiles, supported by targeted notifications (e.g., IVR, SMS), virtual consultations, and CHW-led services tailored to their specific needs.

This pilot demonstrated the feasibility and impact of integrating telemedicine into community health structures to improve maternal and newborn care in underserved settings. The hybrid model combining virtual doctor consultations, CHWs, and SMS-based engagement enhanced access, improved referral completion, and supported positive health-seeking behaviours. Notably, insights from this pilot have already informed early discussions with county health officials regarding the integration of telemedicine into routine MNCH programming. As part of this engagement, stakeholders are exploring a closed-loop solution—which connects virtual care (via the telemedicine provider) with physical care (CHWs and health facilities) through a Health Information Exchange (HIE), ensuring seamless data sharing, case continuity, and patient-centered care across the continuum.

**Figure 7:**
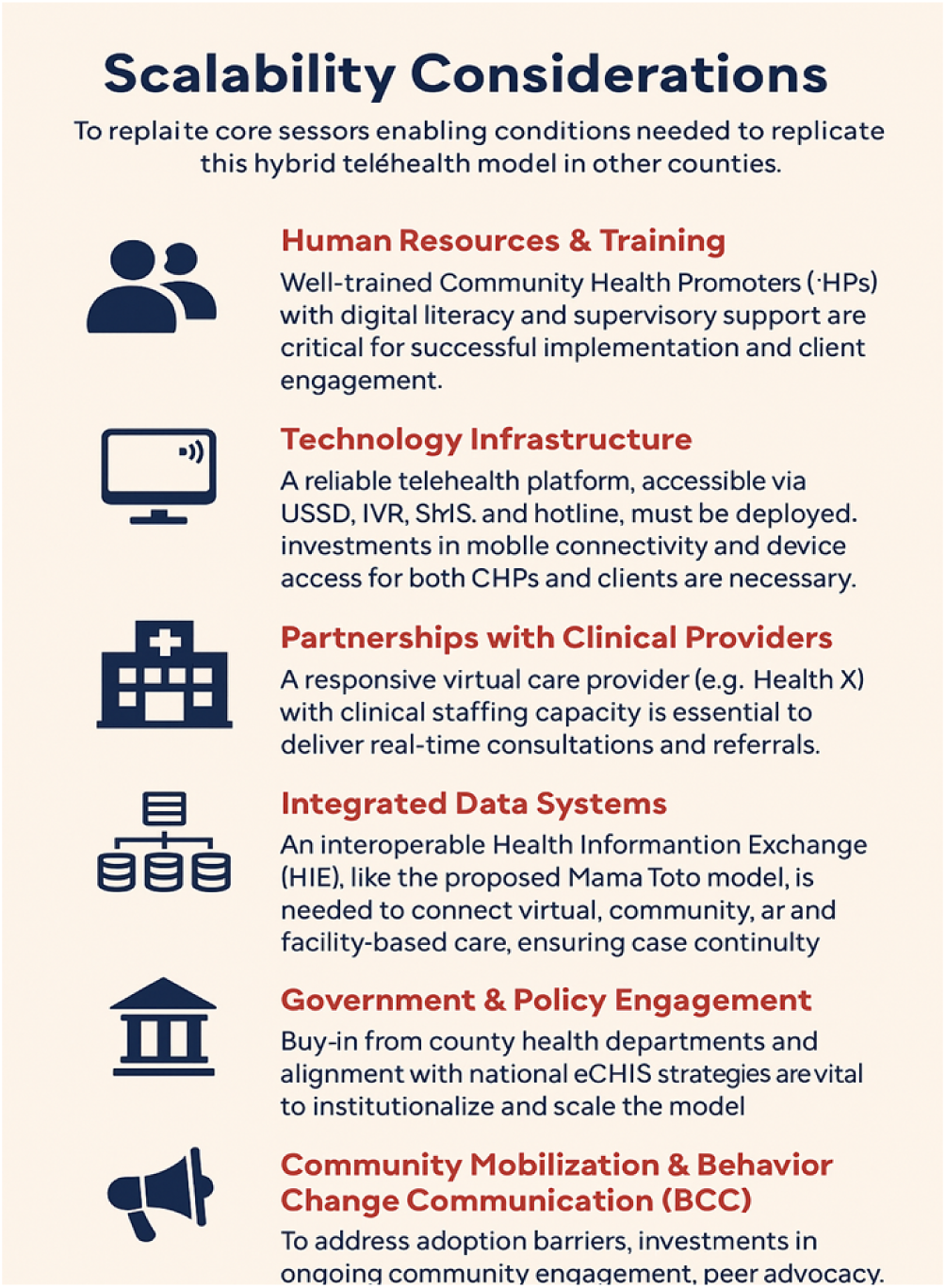
Scalability Considerations

## LIMITATIONS AND FUTURE RESEARCH

While the study provides valuable insights into the potential of telemedicine in community health, it acknowledges certain limitations. Future research should focus on rigorous evaluation methods, including control groups and robust outcome measures, to accurately assess the long-term impact of telemedicine interventions on MNCH. Additionally, further investigations into cost-effectiveness, sustainability, and integration into national health systems will be essential for scaling up telehealth solutions in similar settings.

## CONCLUSION

The ‘*Better Data for Better Decisions’* telemedicine initiative in Kenya demonstrated promising results in enhancing maternal and newborn child health services by integrating technology with the existing community health worker framework. The project exceeded registration targets, and the use of hotline consultations and SMS reminders facilitated increased postnatal care touchpoints and improved appointment adherence. While the study highlights the potential of this hybrid model to strengthen follow-up and referrals, further investigation is needed to quantify the impact on overall healthcare delivery efficiency and long-term policy influence. Insights from this pilot have already informed early discussions with county health officials regarding telemedicine integration into routine MNCH programming.

## Data Availability

Cover letter-Digital health 1.docx The data that support the findings of this study are available from Living Goods, but restrictions apply to the availability of these data due to participant confidentiality. Data are available from the corresponding author upon reasonable request and with permission of Kenyatta University Agriculture and Technology (JKUAT) Ethics Committee.

## Author summary

EA: Conceptualization, methodology, writing, supervision, investigation, project administration, visualization.

TG: Conceptualization, methodology, review and validation, and visualization.

EY: Resources, validation and review

NM: Methodology, writing and validation.

MO: Conceptualization, formal analysis, methodology, supervision.

AK: Review and editing

RO: Methodology, writing and validation.

SS: Conceptualization, methodology, writing, supervision, software and project administration

HA: Conceptualization, methodology, writing, supervision, investigation, resources and project administration

DO: Review and validation

RA: Review and validation

ML: Data curation.

EM: Supervision

GS: Supervision

PA: Supervision

Acknowledge: MoH Busia county, study participants, CHWs, CHAs and research assistants

## Notes

### Competing Interest Statement

The authors have declared no competing interest.

### Funding Statement

The author(s) received no specific funding for this work.

### Author Declarations

Ethical approval was granted by Jomo Kenyatta University Agriculture and Technology (JKUAT) Ethics Committee.

## REFERENCES

1. Cantor AG, Jungbauer RM, Totten AM, Tilden EL, Holmes R, Ahmed A, et al. Telehealth Strategies for the Delivery of Maternal Health Care: A Rapid Review. Ann Intern Med. 2022 Sep;175(9):1285–97.

2. Gajarawala SN, Pelkowski JN. Telehealth Benefits and Barriers. The Journal for Nurse Practitioners. 2021 Feb;17(2):218–21.

3. KNBS and ICF. 2023. Kenya Demographic and Health Survey 2022: Volume 1. Nairobi, Kenya, and Rockville, Maryland, USA: KNBS and ICF.

4. Arias MP, Wang E, Leitner K, Sannah T, Keegan M, Delferro J, et al. The impact on postpartum care by telehealth: a retrospective cohort study. American Journal of Obstetrics & Gynecology MFM. 2022 May;4(3):100611.

5. Hawkins SS. Telehealth in the Prenatal and Postpartum Periods. Journal of Obstetric, Gynecologic & Neonatal Nursing. 2023 Jul;52(4):264–75.

6. Knop MR, Nagashima-Hayashi M, Lin R, Saing CH, Ung M, Oy S, et al. Impact of mHealth interventions on maternal, newborn, and child health from conception to 24 months postpartum in low- and middle-income countries: a systematic review. BMC Med. 2024 May 15;22(1):196.

7. Kachimanga C, Divala TH, Ket JCF, Kulinkina AV, Zaniku HR, Murkherjee J, et al. Adoption of mHealth Technologies by Community Health Workers to Improve the Use of Maternal Health Services in Sub-Saharan Africa: Protocol for a Mixed Method Systematic Review. JMIR Res Protoc. 2023 May 4;12:e44066.

8. Bilal W, Mohanan P, Rahmat ZS, Ahmed Gangat S, Islam Z, Essar MY, et al. Improving access to maternal care in Africa through telemedicine and digital health. Health Planning & Management. 2022 Jul;37(4):2494–500.

9. Atkinson J, Hastie R, Walker S, Lindquist A, Tong S. Telehealth in antenatal care: recent insights and advances. BMC Med. 2023 Aug 30;21(1):332.

9. Cantor AG, Jungbauer RM, Totten AM, Tilden EL, Holmes R, Ahmed A, et al. Telehealth Strategies for the Delivery of Maternal Health Care: A Rapid Review. Ann Intern Med. 2022 Sep;175(9):1285–97.

10. Berkley JA, Walson JL, Bahl R, Rollins N. Differentiating mortality risk of individual infants and children to improve survival: opportunity for impact. The Lancet. 2024 Aug;404(10451):492–4.

